# Myalgic Encephalomyelitis/Chronic Fatigue Syndrome (ME/CFS) is common in post-acute sequelae of SARS-CoV-2 infection (PASC): Results from a post-COVID-19 multidisciplinary clinic

**DOI:** 10.1101/2022.08.03.22278363

**Authors:** H Bonilla, TC Quach, A Tiwari, AE Bonilla, M Miglis, P Yang, L Eggert, H Sharifi, A Horomanski, A Subramanian, L Smirnoff, N Simpson, H Halawi, O Sum-Ping, A Kalinowski, Z Patel, R Shafer, L. Geng

## Abstract

**Background:** The global prevalence of PASC is estimated to be present in 0·43 and based on the WHO estimation of 470 million worldwide COVID-19 infections, corresponds to around 200 million people experiencing long COVID symptoms. Despite this, its clinical features are not well defined.

**Methods:** We collected retrospective data from 140 patients with PASC in a post-COVID-19 clinic on demographics, risk factors, illness severity (graded as one-mild to five-severe), functional status, and 29 symptoms and principal component symptoms cluster analysis. The Institute of Medicine (IOM) 2015 criteria were used to determine the ME/CFS phenotype.

**Findings:** The median age was 47 years, 59·0% were female; 49·3% White, 17·2% Hispanic, 14·9% Asian, and 6·7% Black. Only 12·7% required hospitalization. Seventy-two (53·5%) patients had no known comorbid conditions. Forty-five (33·9%) were significantly debilitated. The median duration of symptoms was 285·5 days, and the number of symptoms was 12. The most common symptoms were fatigue (86·5%), post-exertional malaise (82·8%), brain fog (81·2%), unrefreshing sleep (76·7%), and lethargy (74·6%). Forty-three percent fit the criteria for ME/CFS.

**Interpretations:** Most PASC patients evaluated at our clinic had no comorbid condition and were not hospitalized for acute COVID-19. One-third of patients experienced a severe decline in their functional status. About 43% had the ME/CFS subtype.

**Funding:** The study did not received funding.

## Introduction

While most patients recover within weeks of SARS-CoV-2 infection, others experience debilitating symptoms that persist beyond the acute period (1). The overall global prevalence of post-COVID-19 conditions is estimated at 0·43 of acute cases (hospitalized 0·54 and non-hospitalized 0·36)(2). These post-COVID conditions, collectively known as a Post-Acute Sequelae of SARS-CoV-2 infection (PASC), or long COVID, are increasingly recognized even in patients who experience asymptomatic or mild SARS-CoV-2 infection(3). The Center for Disease Control and Prevention (CDC) defines post-COVID conditions as symptoms persisting beyond 28 days after infection(4), while the U.K. National Institute for Health and Care Excellence (NICE)(5) and the World Health Organization (WHO) define this syndrome as symptoms persisting beyond 12 weeks after infection(6). The American Academic of Physical Medicine and Rehabilitation (AAPMR&R) estimates that there are more than 24 million cases of long COVID as of May, 2022 (7).

PASC is most often characterized by extreme fatigue exacerbated by exertion, referred to as post-exertional malaise, difficulty with concentration and memory often referred to as brain fog, sleep disturbances, headaches, chest pain, and shortness of breath(2, 8). PASC can range from mild to severe and incapacitating, interfering with patients’ daily activities and work requirements(2, 8).

In an online, multi-national survey of 3,762 participants with confirmed or suspected COVID-19, fatigue, post-exertional malaise, and brain fog were the most frequently reported symptoms six months after SARS-CoV-2 infection(8). This cluster of symptoms shares similar features with ME/CFS, an often-debilitating disease that has a worldwide prevalence close to one percent, is also believed to frequently arise following a viral infection(9, 10). Women are affected 1·5 to 2 times more often than men(2). In two small cohorts of 41 and 42 COVID-19 patients approximately 45% met the ME/CFS diagnostic criteria(11, 12). The pathobiological mechanism of ME/CFS is not known, but the large number of PASC patients presenting with features of this syndrome may allow us to better understand both conditions. The goals of our study are two-fold: 1. Characterize our clinical cohort in terms of demographics and clinical presentations. 2. Estimate the prevalence of ME/CFS phenotype in PASC based on the IOM ME/CFS criteria(9).

## Methods

### Study design and participants

The Stanford referral Post-Acute COVID-19 Syndrome (PACS) Clinic is a multidisciplinary center designed to provide clinical expertise in post-COVID-19 conditions, standardization of data collection and clinical management, and integration of research efforts. We standardized the clinical assessment by developing a clinic template embedded in the Electronic Health Records (EHR), allowing the extraction of the same data retrospectively. We used Research Electronic Data Capture (REDCap) and Microsoft Excel platforms for data collection and analysis. The study was approved by the Stanford University Institutional Review Board.

One hundred-forty consecutive adult patients with a history of COVID-19 were seen in the Stanford PACS clinic between May 18, 2021, and February 1, 2022. Referral criteria included a history of symptomatic SARS-CoV-2 infection, a diagnostic test for SARS-CoV-2 by either PCR, antigen detection, or a positive serology before SARS-CoV-2 vaccination, and persistent symptoms for at least 28 days following infection. In addition, information about persistent symptoms from each patient was obtained from questionnaire seven days before the clinic visit and from the patient’s EHR. The questionnaire includes: (i) the 29 symptoms reported to occur commonly in patients with acute COVID-19 seven days before the clinic visit; (ii) the severity of each symptom based on the Likert scale (1 mild symptom and 5 severe) (iii) SARS-CoV-2 vaccination status; (iv) the Post COVID-19 Functional Status Scale (FSS) which classifies patients as either asymptomatic (level 1), symptomatic without limitations (level 2), symptomatic with reduced daily activity (level 3), symptomatic with a struggle to perform daily activities (level 5), or incapacitated and bedridden (level 5) (2, 13). From the EHR, we extracted the following: (1) demographics including age, sex, and self-identified race/ethnicity; (2) laboratory results and radiological data obtained before the initial office visit; and (3) vital signs, body mass index (BMI), oxygen saturation, orthostatic blood pressure, heart rate measurements obtained at the initial visit.

An identical questionnaire was sent to each patient prior to their scheduled clinic visit, collecting the same information as we did for the acute infection (Supplemental material). For patients whose initial clinic visit was six months or more after their infection, we analyzed the symptoms to determine whether they met the Institute of Medicine (IOM) 2015 diagnostic criteria for ME/CFS(9). Patients with a history of ME/CFS or other condition that explains the fatigue symptom prior to COVID-19 were excluded from this analysis.

### Statistical analysis

To explore how the symptoms correlated with each other in our study cohort, we utilized R (version 3·6·1) cluster analysis algorithms in 134 post-COVID-19 patients based on principal components analysis (PCA) to visualize the distribution of symptoms(14, 15). We used the Wilcoxon Rank Sum Test to calculate the difference in the number of symptoms between males and females, Chi-square to calculate the difference in symptoms between male and female and the Pearson Coefficient Correlation (r) to compare the frequency of the symptoms and severity.

## Results

Among the 140 patients referred to the Stanford PASC clinic, six patients were excluded because of lack of a diagnostic test for SARS-CoV-2 infection or an incomplete questionnaire. For the remaining 134 patients, the median age was 47 years old (IQR: 30-60; range: 20 to 88). Seventy-nine (59%) were female, 66 self-identified as White (49·3%), 23 as Hispanic (17·2%), 20 as Asian (14·9%), and 9 as Black (6·7%). Seventeen (12·7%) required hospitalization for their acute infection and were considered to have had severe disease. Two of these patients required Intensive Care Unit (ICU) admission. Sixty-two (46·3%) patients had a comorbid condition known to predispose to severe disease including obesity (BMI > 30 kg/m^2^) 47 (35·1%), hypertension 24 (17·9%), chronic lung disease 16 (11·9%), diabetes 9 (6·7%), immunosuppressive therapy 5 (3.7%) and/or cardiovascular disease 4 (3%). One hundred and nine (82%) patients had some degree of functional limitation (i.e., FSS of 3, 4, or 5), including 45 (33·9%) who exhibited significantly compromised wellbeing (FSS of 4 or 5) (Table 1).

**Table 1.** Patient’s characteristics and distribution PASC cohort and Chronic Fatigue Cohort.

| **Characteristics** | **PASC Cohort**<br>**(N =134)** | **Chronic Fatigue Cohort**<br>**(N=105) Persistent symptoms over six months** |  |  |
| --- | --- | --- | --- | --- |
|  |  | **ME/CFS** | **No ME/CFS** | **Total** |
| Age (years) Median (Range) | 47(20–88) | 50(23-75) | 47(21-88) | 47(21-88) |
| Sex: Female/Male (Female%) | 79/55 (59%) | 26/19 (58%) | 42/18(70%) | 68/38 |
| Days Post-Covid Median (Range) | 285.5 (34-792) | 318(183-686) | 310(149-792) | 310(183-792) |
| **Diagnostic Test N (%)** |  |  |  |  |
| Molecular/PCR | 119(88.8%) | 38(84%) | 54(90%) | 92 |
| Antigen | 5(3.7%) | 2(6%) | 3(5%) | 5 |
| Antibodies (+) before Vaccination | 10(7.5%) | 5(10%) | 3(5%) | 8 |
| **Treatment** |  |  |  |  |
| Ambulatory | 117/134(87.3%) | 44(85%) | 57 (95%) | 101 |
| Hospital /ICU | 17/134(12.7%) | 1(2%)/0 | 3(5%)/1 | 4/1 |
| **Functional Status (%)** |  |  |  |  |
| V (%) | 9(6.8%) | 5(11%) | 3(5%) | 8 |
| IV (%) | 36 (27.1%) | 19 (42%) | 13(22%) | 32 |
| III (%) | 64(48.1%) | 20(44%) | 27(45%) | 47 |
| I-II (%) | 20(15%) | 1(3%) | 17(28%) | 18 |
| **Co-morbid Condition N (%)** |  |  |  |  |
| None | 72(53.7%) | 17(38%) | 36(60%) | 53 |
| One | 37(27.6%) | 13(29%) | 18(30%) | 31 |
| Two | 15(11.2%) | 7(15%) | 5(8%) | 12 |
| Three | 7(5.2%) | 9(18%) | 1(2%) | 10 |
| **BMI kgm<sup>2</sup> (median, range)** |  |  |  |  |
| All | 26.2(16.9–46.6) | 29.23(18.9-47.9) | 25.6(18.3-41.1) | 27.2(16.9-46.6) |
| ≥30 (%) | 31(33.6%) | 22(49%) | 17(28%) | 39 |
| ≥35 (%) | 21(15%) | 10(22%) | 8(13%) | 19 |
| **Race/ethnicity** |  |  |  |  |
| White | 66(49.3%) | 17(38%) | 34(57%) | 51 |
| Hispanic | 23(17.2%) | 13(28%) | 7(12%) | 20 |
| Black | 9(6.7%) | 2(4%) | 4(7%) | 6 |
| Asian | 20(14.9%) | 4 (8%) | 8(13%) | 12 |
| Others | 2(1.4%) | 1(2%) | 1(1%) | 2 |
| No Data | 15(11%) | 8(18%) | 6(10%) | 14 |
| **TOTAL** | 134(100%) | 45(43%) | 60(57%) | 105(100%) |

PASC: Post-Acute Sequela of SARS-Cov-2 infection. ME/CFS: Myalgic Encephalomyelitis/Chronic Fatigue Syndrome

The duration of the symptoms at the time of the initial office visit ranged from 34 to 792 days with a median duration of 285·5 days. Of the 29 symptoms assessed in the study, the median number per patient was 12 (range: 1 to 25 symptoms). The most common symptoms were fatigue (86·5%), post-exertional malaise (82·8%), brain fog (81·2%), unrefreshing sleep (76·7%), and daytime sleepiness (74·6%). The median number of symptoms in female patients was greater than the median number in male patients (13 vs. 10, p=0·005). The most common symptoms significantly higher in the female population were fatigue, insomnia, and change in taste or dysgeusia (P values = 0·001, 0·044, and 0·01 respectively). There was a significant correlation between the frequency and the severity of the symptom (Likert scale 4 and 5) (Table 2, Figure. 3) (Correlation 0·9209994, P<0·0001).

**Table 2.** Distribution rates of the most common 15 symptoms in PASC patients by sex and severity.

| Symptom | Rates by Sex (%) |  | P Value | Rates Likert Scale 4-5** | Total (%)** |
| --- | --- | --- | --- | --- | --- |
|  | Female | Male |  |  |  |
| Fatigue | 88.6 | 81.8 | 0.0011* | 61.8 | 86.5 |
| Post Exertional Malaise | 84.8 | 80 | 0.4675 | 66.6 | 82.8 |
| Brain Fog | 81 | 80 | 0.8840 | 50 | 81.2 |
| Unrefreshing Sleep | 81 | 69.1 | 0.1113 | 49 | 76.7 |
| Lethargic | 78.5 | 69.1 | 0.2191 | 57 | 74.6 |
| Insomnia | 74.7 | 58.2 | 0.0441* | 44 | 65.9 |
| Headache | 65.8 | 63.6 | 0.9129 | 31.4 | 62.1 |
| Anxiety/Depression | 57 | 67.3 | 0.3226 | 39.5 | 52.6 |
| Lightheadedness | 57 | 45.5 | 0.1895 | 32.8 | 50.8 |
| Gastrointestinal | 57 | 40 | 0.1289 | 20.9 | 48.9 |
| Shortness of Breath | 51.9 | 41.8 | 0.3145 | 30.1 | 43.2 |
| Nasal Congestion | 49.4 | 32.7 | 0.07590 | 21.4 | 42 |
| Changes of Smell | 49.4 | 32.7 | 0.1024 | 32.7 | 41.2 |
| Changes of Taste | 46.8 | 27.3 | 0.01* | 31.5 | 39.2 |
| Cough | 44.3 | 29.1 | 0.074 | 23.5 | 36.6 |

\*P value < 0.05 Chi-square
\*\*Pearson correlation coefficient 0.92, P value <0.0001

In the PCA analysis, the direction of arrows in the correlation circle illustrates the extent to which symptoms were likely to occur in the same patient while the factor map indicates the contribution of different symptoms to the first five components (Figure. 2). The analysis suggests that fatigue, PEM, daytime sleepiness (lethargy), brain fog, and unrefreshing sleep were likely to occur together. These symptoms were major contributors to the first component while anosmia and nasal congestion were major contributors to the second component.

**Figure 1.**
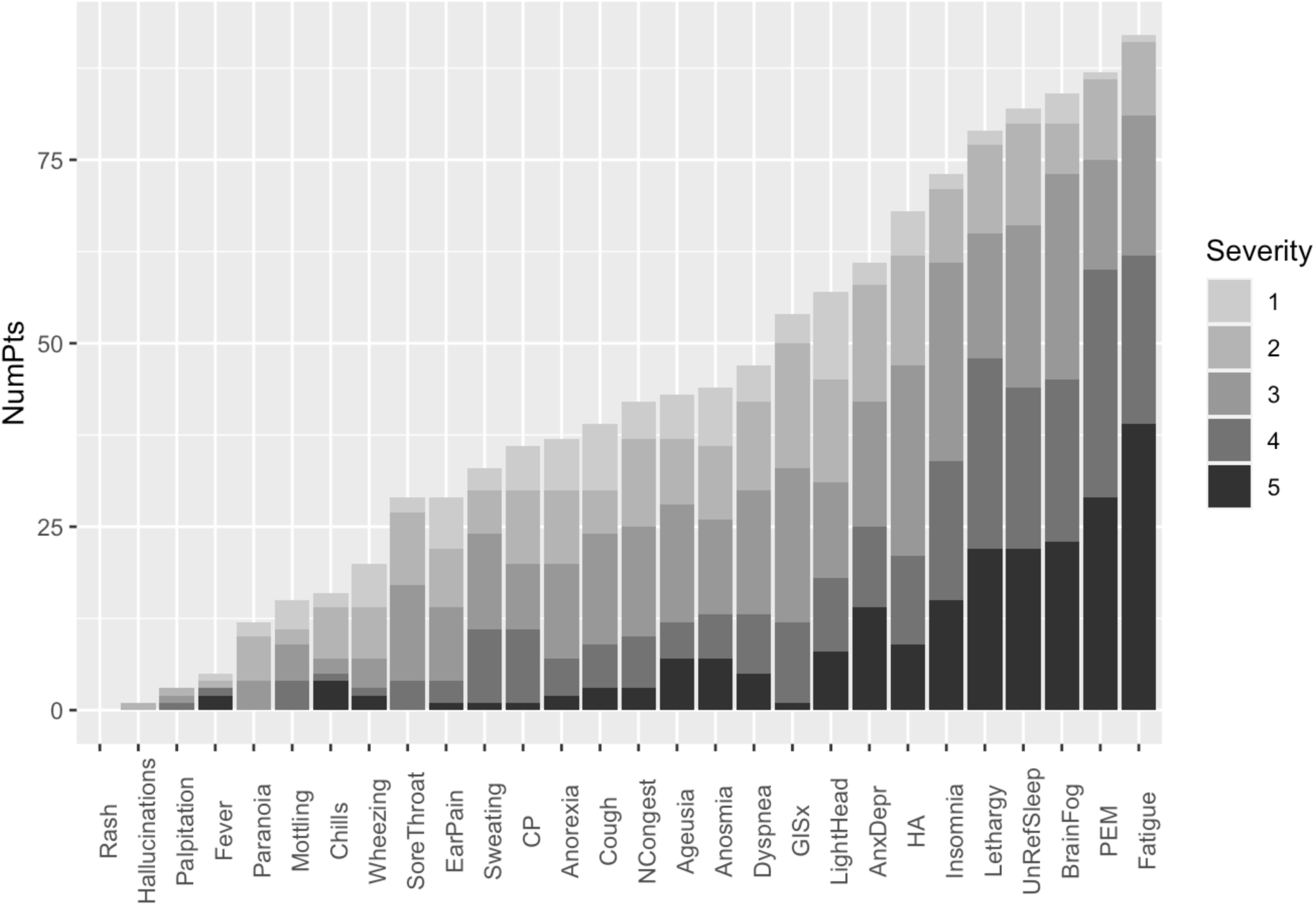
Distribution of the number and severity of the symptoms after six months of the SARS-Cov-2 Infection. The darker color represents a more severe is the symptom. CP: Chest pain; GISx: Gastrointestinal symptoms; Anex/Depr: Anxiety/Depression; HA: Headache; Lethargy: daytime sleepiness; UnRefrSleep: Unrefreshed sleep; PEM: Post-Exertional Malaise. Distribution of the frequency and severity of the symptoms on the PASC clinic intake questionnaire for subgroup of 105 patients with persistent symptoms for six or more months. Symptom severity was measured on the Likert scale with a severity score of 5 being the most severe. Abbreviations:

**Figure 2.**
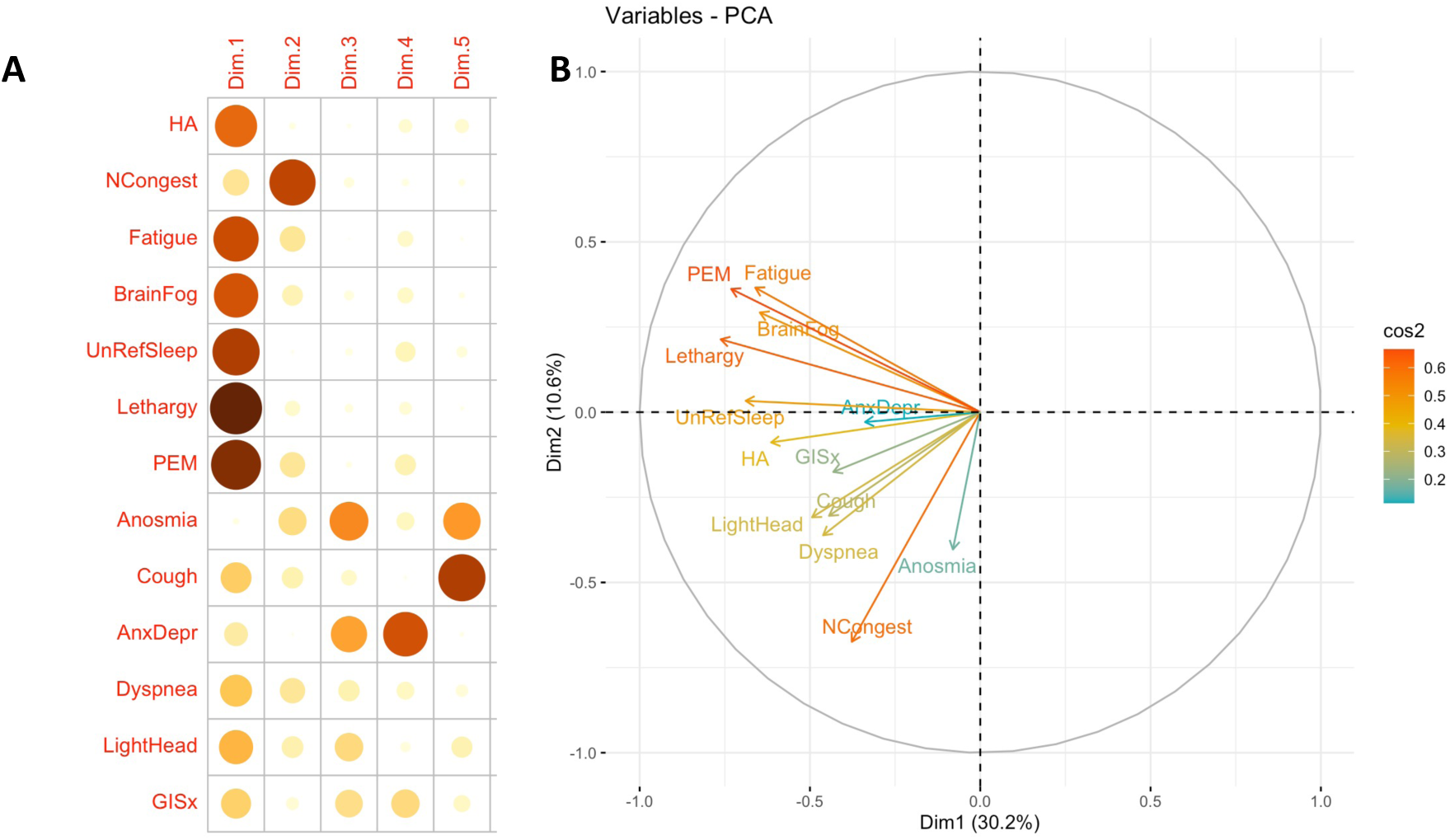
Principal components analysis of PASC symptoms, mapped to the first five dimensions and circle correlation. Quality of representation of 13 most common symptoms mapped to the first five dimensions (A) and principal components analysis correlation circle (B). Insomnia and ageusia were not included in the analysis. The closer the variable to the correlation circle, the better the representation on the factor map. The quality of the representation of the symptom on the first five dimensions is measured by the squared cosine between the symptom vector and its projection on the dimension. The proportions on the left side of the factor map represent a color scale. Reference: (10.18637/jss.v025.i01) Abbreviations:

Because fatigue was the most common and severe of all symptoms, we decided to explore the prevalence of the ME/CFS phenotype in our study population. One hundred and five patients (69%) had persistent symptoms longer than six months and the top six most common and severe symptoms were fatigue, post-exertional malaise (PEM), brain fog, unrefreshing sleep, daytime sleepiness, and insomnia (Figure 2). From the 48 (46%) of patients who fit the IOM criteria for ME/CFS (A substantial reduction or impairment in the ability in the pre-illness activities, PEM, unrefreshing sleep, and one of the two following cognitive impairment, or orthostatic intolerance), 3 were excluded from the ME/CFS cohort because of the following reasons: 1 patient with history of rheumatoid arthritis/Lupus/Sjogren’s, MDS and asthma, 1 with history of fibromyalgia, and 1 with history of Scleroderma/ME/CFS. Therefore, 45 (43%) of the study cohort fulfill the criteria for ME/CFS. Similar to the PASC cohort, the ME/CFS cohort was predominantly female, non-hospitalized, and healthy, and obesity was the most common risk factor (BMI > 30 Kg/m^2^), 22 (49%). Furthermore, 53% of ME/CFS had significant decline in their functionality (FSS 4 and 5) (Table.1).

## Discussion

This manuscript presents the characteristics of 134 patients referred to a long COVID clinic who had who have history of SARS-CoV-2 infection and more than 28 more days of symptoms. The median age in our cohort was 47 years and ranged from 20 to 88 years, with a female predominance of 59%. Similar age and sex distribution was reported in several other studies(2, 16). In contrast to hospitalized patients, females PASC were represented in a lower proportion, less severe symptoms, and lower mortality than males(17-19). These differences in sex distribution could be explained by the variations in the immune response between males and females, and the fact that female patients have more robust inflammatory, antiviral, and humoral immune responses, biological differences on sex hormones, and expression and regulation of angiotensin-converting enzyme 2 (ACE2)(18, 20). Our study cohort, similar to other PASC studies, the subjects were predominantly white females, with obesity(17, 21, 22) characteristics associated with lower likelihood for full recovery(23). In contrast, some racial and ethnic minority groups, such as Native American Indians, Alaska Natives, Hispanic and Black, have been shown to have a disproportionately higher risk for infection, severity of illness, hospitalization, and deaths. Those groups were underrepresented in our study(24).

The large majority (87%) of our PASC patients had not been hospitalized or required oxygen for COVID-19 disease. The prevalence of PASC in asymptomatic patients with the mild disease is reported between 30-60% (3, 25). We can postulate that unknown factors besides hypoxemia and hospitalization are the drivers of PASC symptoms such as virus persistence, overactivation of the immune system, amyloid fibrin microclots, auto-antibodies, virus reactivation and others, may play a role in the pathobiology of this illness(26).

In our study, the duration of symptoms ranged from 34-792 days, with a median duration of 285·5 days. Females experienced more symptoms than males. The most common symptoms were fatigue, post-exertional malaise, brain fog, unrefreshing sleep, and daytime sleepiness (Supplement Table. 3). This is similar to a meta-analysis which found that fatigue/weakness, myalgia/arthralgia, depression, anxiety, memory loss, concentration difficulties, dyspnea, and insomnia, were the most prevalent symptoms(2, 27). We observed a significant correlation between frequency with severity of the symptoms and the ME/CFS symptoms clustered with one another. Fatigue was the most prevalent symptom across the PASC studies(2, 27). Post-viral fatigue was commonly reported after a viral infection such as Influenza, SARS/MERS, and Ebola(28). In a study on 233 SARS survivors, 40·3 % reported having chronic fatigue, and 27·1% met the criteria for ME/CFS; after influenza with H1N1, ME/CFS has reported 2·08 cases/100,000 person-month(28). In our study, 43% of the selected cohort fulfilled all the ME/CFS criteria, which is similar to 45% reported by Mancini et al and Cedor et al.(11, 12). Like ME/CFS, the ME/CFS-PASC phenotype was more prevalent among the non-hospitalized female population (Table. 1). The clinical similarities in our study cohort between ME/CFS and ME/CFS-PASC allow us to suggest common pathobiology. Those similarities include a preceding a viral illness(29), increase in inflammatory cytokines, neuroinflammatory change, mitochondria dysfunction, and alteration in NK cell function(30).

Our study has several limitations. First, this study only represents the experience of a single center in Northern California located in a generally affluent area with a bias toward specific populations. The selection of this cohort may skew our population in the follow ways: 1) this is a referred population with multiple and more severe symptoms 2) our clinics have a lower proportion of underrepresented minority populations. Therefore, a multicenter study that includes a more diverse and larger population is necessary to corroborate our findings.

In summary, about half of the PASC patients with more than six months of symptoms fulfilled the ME/CFS criteria; the majority of patients seen in our PACS clinic were female, with a median age of 47 years old, with obesity as the most common comorbidity. The majority of the patients had mild to moderate acute infection and were healthy prior to their COVID infection. Fatigue, post-exertional malaise, brain fog, unrefreshing sleep, and daytime sleepiness were the most prevalent and severe symptoms. This commonality between ME/CFS and ME/CFS-PASC may suggest a shared pathobiology. Therefore, defining specific subtypes within the umbrella of PASC/post-COVID conditions can help us understand different pathogenic mechanisms to tailor treatment.

## Supporting information

Questionnaries and figures

## Data Availability

All data produced in the present study are available upon reasonable request to the authors.

## Data sharing

The data collected in the study will be available in the supplementary files and additional dataset for this manuscript are also available from the corresponding author upon request.

## Acknowledgements

Stanford Health Care, and Stanford Department of Medicine for the clinic support.

## Authors contributions

**Hector Bonilla:** Corresponding author, conception and design of the study, data collection, analysis, draft, editing of the manuscript, full access full access to the whole data, to the whole data, final version approval.

**Quach TC:** data collection, full access to the whole data, analysis, draft, and editing of the manuscript.

**Tiwari A:** data collection, full access to the whole data, analysis, draft, and editing of the manuscript.

**Bonilla AE:** data collection, full access to the whole data, analysis, draft, and editing of the manuscript.

**Miglis M:** conception and design of the study, full access to the whole data, and editing of the manuscript.

**Yang P:** conception and design of the study, full access to the whole data, and editing of the manuscript.

**Eggert L:** conception and design of the study, full access to the whole data, and editing of the manuscript.

**Sharifi H:** conception and design of the study full access to the whole data and editing of the manuscript.

**Horomanski A:** conception and design of the study full access to the whole data and editing of the manuscript.

**Subramanian A:** conception and design of the study, full access to the whole data, and editing of the manuscript.

**Smirnoff L:** conception and design of the study, full access to the whole data, and editing of the manuscript.

**Simpson N:** conception and design of the study, full access to the whole data, and editing of the manuscript.

**Halawi H:** conception and design of the study, full access to the whole data, and editing of the manuscript.

**Sum-Ping O:** conception and design of the study full access to the whole data and editing of the manuscript.

**Kalinowski A:** conception and design of the study full access to the whole data and editing of the manuscript.

**Patel Z:** conception and design of the study, full access to the whole data, and editing of the manuscript.

**Shafer R:** conception and design of the study, full access to the whole data, data collection, analysis, draft, editing of the manuscript, final version approval.

**Geng L:** conception and design of the study, full access to the whole data, data collection, analysis, draft, editing of the manuscript, final version approval.

