## Supplementary material for "Myalgic Encephalomyelitis/Chronic Fatigue Syndrome (ME/CFS) is common in post-acute sequelae of SARS-CoV-2 infection (PASC): Results from a post-COVID-19 multidisciplinary clinic": Questionnaries and figures

Stanford
HEALTH CARE
STANFORD MEDICINE

### First assessment Post Covid infection

Name: ___________________________________ Date of Birth: _________

COVID-19 Symptoms: Start date _____________ Duration of Symptoms (days)__________

Date of Positive Test *(if not done at Stanford, please provide copy of the results)*: _____________

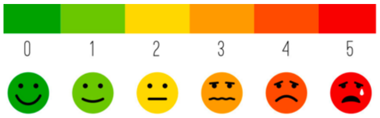

**What were your symptoms during the initial COVID-19 infection? Please also select severity.**

| **Symptoms during acute COVID-19 Infection** | Yes | No | **Severity (1=mild, 5=severe)** |
|---|---|---|---|
| Fever | | | |
| Chills | | | |
| Headache | | | |
| Decrease appetite | | | |
| Nose congestion | | | |
| Sore throat | | | |
| Fatigue | | | |
| Brain fog or confusion | | | |
| Unrefreshing sleep | | | |
| Difficulty sleeping | | | |
| Daytime sleepiness | | | |
| More fatigue with activity | | | |
| Change in smell | | | |
| Change in taste | | | |
| Ear pain | | | |
| New anxiety or depression | | | |
| Paranoid thoughts | | | |
| Hallucinations | | | |
| Cough | | | |
| Chest pain | | | |
| Difficulty breathing at rest | | | |

| |
|---|
| Difficulty breathing while walking |
| Wheezing |

| |
|---|
| Lightheadedness on standing |
| Fainting spells |
| Changes in sweating (more or less) |
| Nausea, vomiting, or diarrhea, bloating or constipation |
| Changes in color of hands or feet |
| Urinary difficulties |

*modified scale from reference: Klok FA, Boon GJAM, Barco S, et al. The Post-COVID-19 Functional Status scale: a tool to measure functional status over time after COVID-19. Eur Respir J 2020; 56: 2001494

### New Patient Questionnaire – Post-COVID Infection

(Please complete within 7 days of the office visit)

Name: ________________________________ Date of Birth: _______________

COVID-19 Symptoms: Start date ____________ Duration of Symptoms (days): __________

Date of Positive Test *(if not done at Stanford, please provide copy of the results)*: ________________

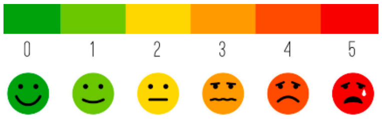

**What symptoms are you currently experiencing? Select severity from scale of 1 to 5.**

| **Current symptoms** | Yes | No | Severity (1=mild, 5=severe) |
|---|---|---|---|
| Fever | | | |
| Chills | | | |
| Headache | | | |
| Decrease appetite | | | |
| Nose congestion | | | |
| Sore throat | | | |
| Fatigue | | | |
| Brain fog or confusion | | | |
| Unrefreshing sleep | | | |
| Difficulty sleeping | | | |
| Daytime sleepiness | | | |
| More fatigue with activity | | | |
| Change in smell | | | |
| Change in taste | | | |
| Ear pain | | | |
| New anxiety or depression | | | |
| Paranoid thoughts | | | |
| Hallucinations | | | |
| Cough | | | |
| Chest pain | | | |
| Difficulty breathing at rest | | | |
| Difficulty breathing while walking | | | |
| Wheezing | | | |
| Lightheadedness on standing | | | |
| Fainting spells | | | |
| Changes in sweating (more or less) | | | |
| Nausea, vomiting, diarrhea, bloating or constipation | | | |
| Changes in color of hands or feet | | | |
| Urinary difficulties | | | |

**What is your current functional status in this post-acute COVID-19 phase?**

| Current Functional Status | Yes | No | Stage |
| --- | --- | --- | --- |
| No symptoms | | | I |
| No limitation but I feel some symptoms | | | II |
| I avoid some of my daily activities | | | III |
| I struggle to take care of myself | | | IV |
| I am in bed all or nearly all the time | | | V |
| I was hospitalized for COVID-related symptoms | | | Severe |

*

**Have you been vaccinated for COVID-19?**

| Vaccine Type | Select one below | Date, first dose | Date, second dose |
| --- | --- | --- | --- |
| BioNTech, Pfizer | | | |
| Moderna, NIAID | | | |
| Johnson & Johnson (JJ) | | | |
| No vaccination | | | |

*modified scale from reference: Klok FA, Boon GJAM, Barco S, et al. The Post-COVID-19 Functional Status scale: a tool to measure functional status over time after COVID-19. Eur Respir J 2020; 56: 2001494

Supplementary Tables (2 and 3) **Supplementary Tables.**

**Table 3. Distribution of Comorbid Condition on PASC Population.**

| **Comorbidities** | **Number/Percentage** |
| --- | --- |
| BMI $\geq$ 30 | 47 (35.1%) |
| Hypertension | 24 (17.9%) |
| Chronic Lung Disease | 17 (12.7%) |
| Diabetes Mellitus | 9 (6.7%) |
| Cardiovascular Disease | 4 (3%) |
| Immunosuppressive therapy | 5 (3.7%) |
| HIV (+) | 1 (0.7%) |
| Cirrhosis | 1 (0.7%) |

PASC: Post-Acute Sequela of SARS-Cov-2 infection

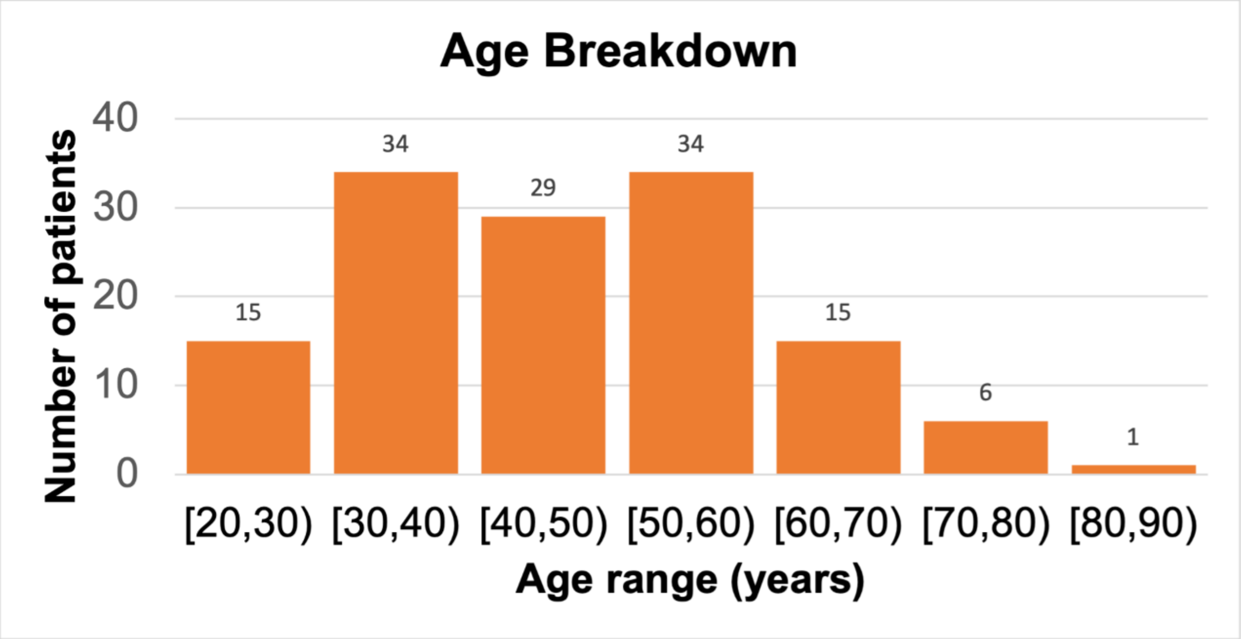
Age Breakdown
Number of patients
40
30
20
10
0
15
34
29
34
15
6
1
[20,30)
[30,40)
[40,50)
[50,60)
[60,70)
[70,80)
[80,90)
Age range (years)

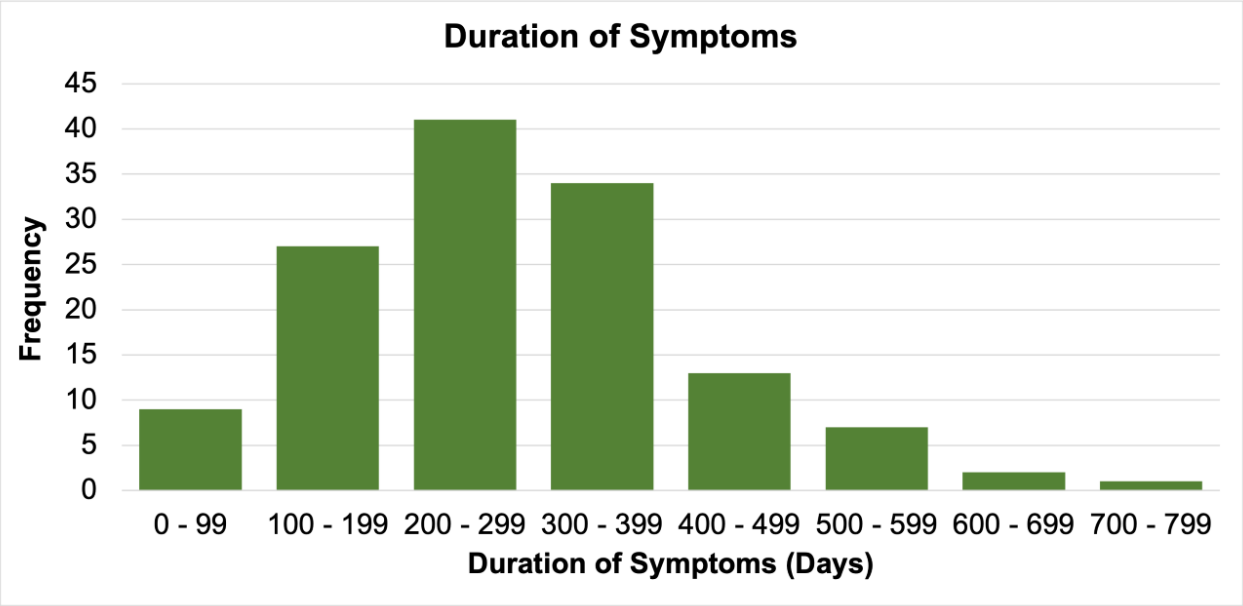
Duration of Symptoms
Frequency
45
40
35
30
25
20
15
10
5
0
0 - 99
100 - 199
200 - 299
300 - 399
400 - 499
500 - 599
600 - 699
700 - 799
Duration of Symptoms (Days)

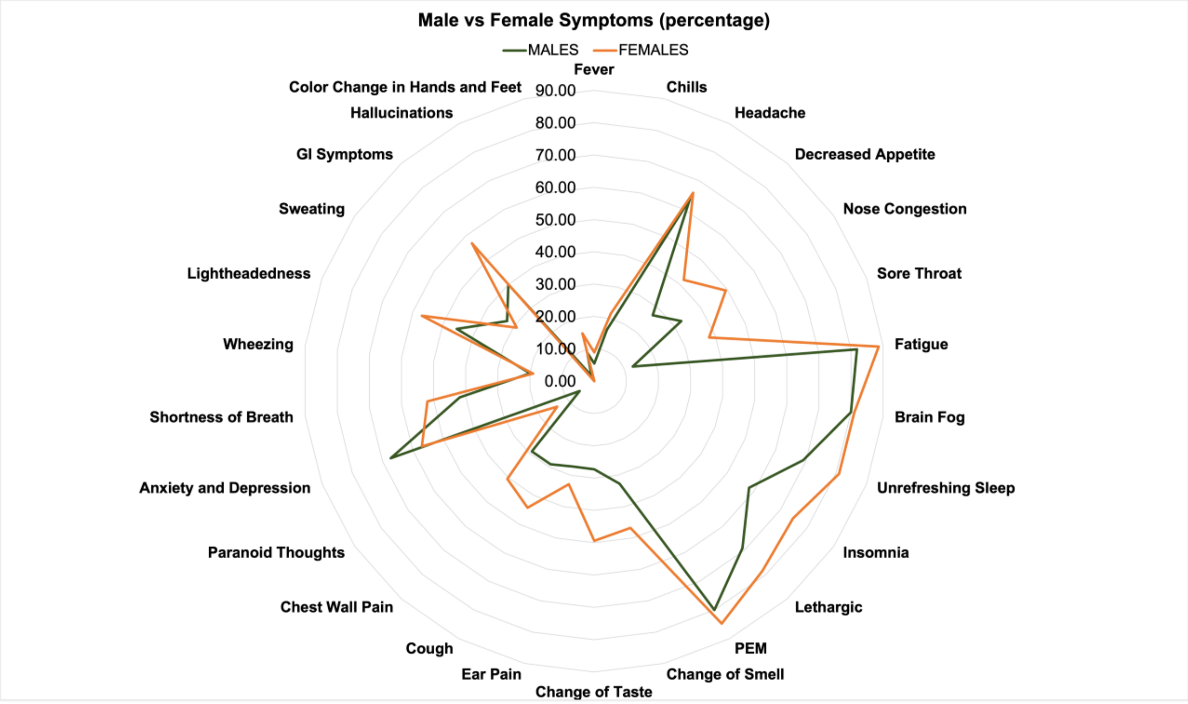

Male vs Female Symptoms (percentage)
MALES
FEMALES
Fever
Chills
Headache
Decreased Appetite
Nose Congestion
Sore Throat
Fatigue
Brain Fog
Unrefreshing Sleep
Insomnia
Lethargic
PEM
Change of Smell
Change of Taste
Ear Pain
Cough
Chest Wall Pain
Paranoid Thoughts
Anxiety and Depression
Shortness of Breath
Wheezing
Lightheadedness
Sweating
GI Symptoms
Hallucinations
Color Change in Hands and Feet
90.00
80.00
70.00
60.00
50.00
40.00
30.00
20.00
10.00
0.00

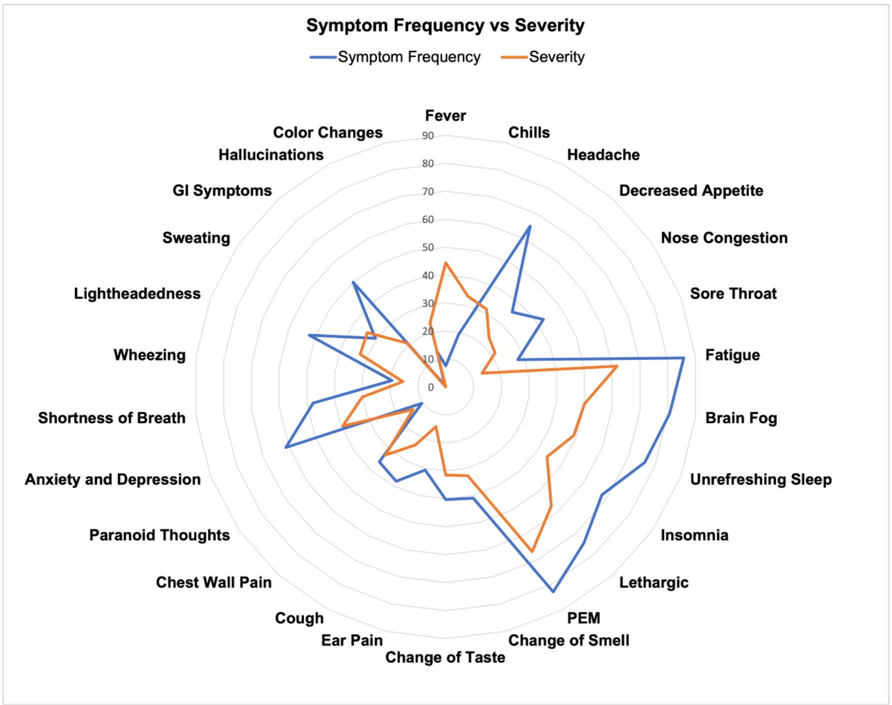

Symptom Frequency vs Severity
Symptom Frequency
Severity
Fever
Chills
Headache
Decreased Appetite
Nose Congestion
Sore Throat
Fatigue
Brain Fog
Unrefreshing Sleep
Insomnia
Lethargic
PEM
Change of Smell
Change of Taste
Ear Pain
Cough
Chest Wall Pain
Paranoid Thoughts
Anxiety and Depression
Shortness of Breath
Wheezing
Lightheadedness
Sweating
GI Symptoms
Hallucinations
Color Changes

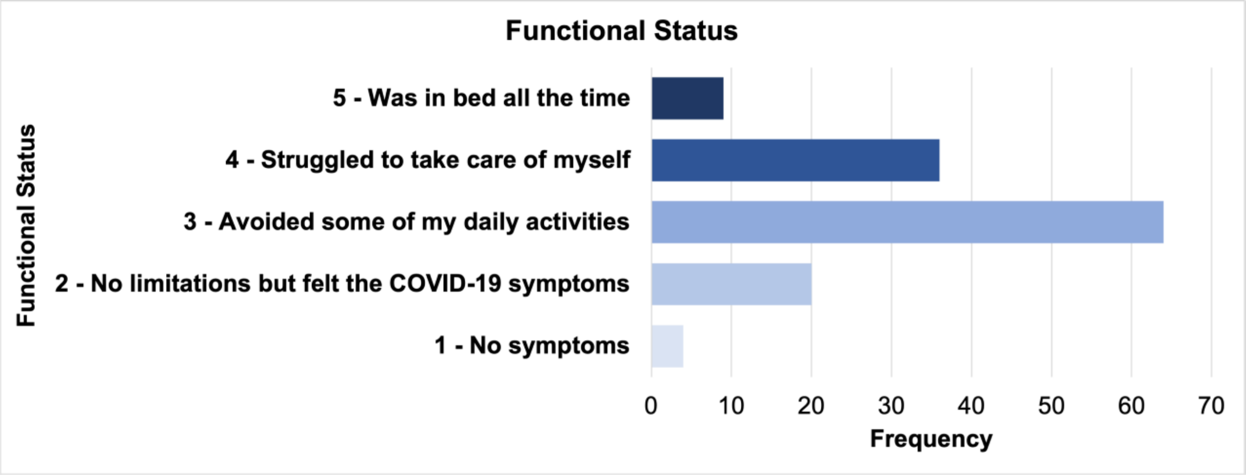
Functional Status
5 - Was in bed all the time
4 - Struggled to take care of myself
3 - Avoided some of my daily activities
2 - No limitations but felt the COVID-19 symptoms
1 - No symptoms
Functional Status
0
10
20
30
40
50
60
70
Frequency

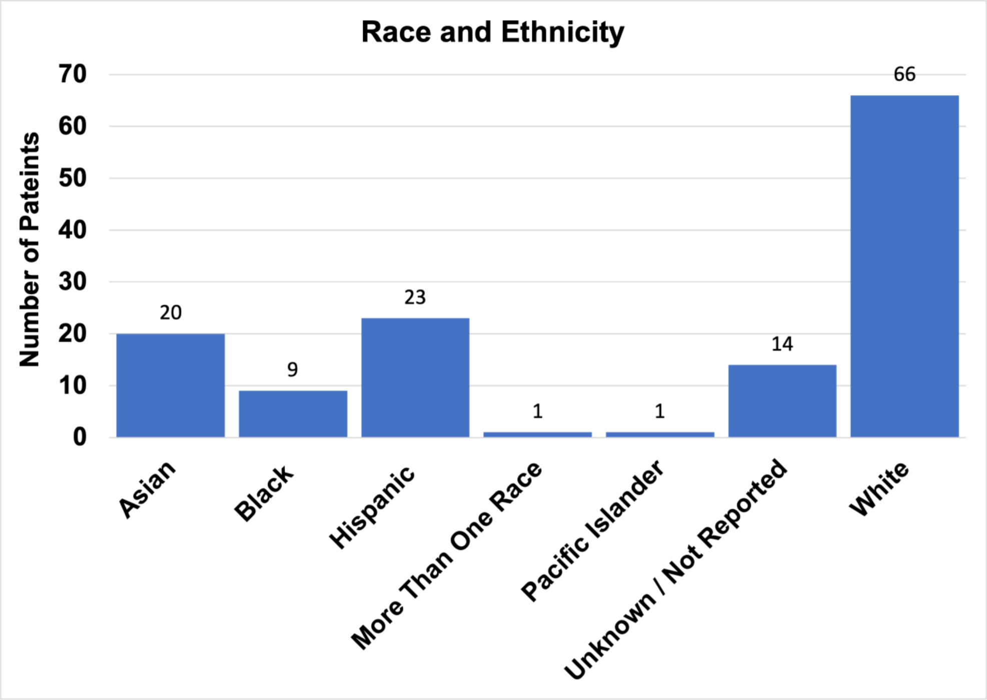
Race and Ethnicity
Number of Pateints
70
60
50
40
30
20
10
0
20
9
23
1
1
14
66
Asian
Black
Hispanic
More Than One Race
Pacific Islander
Unknown / Not Reported
White

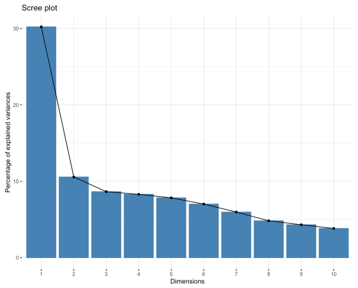

#### 13 most common symptoms

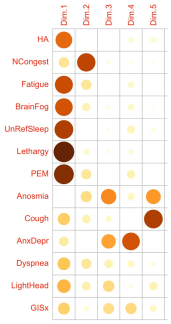

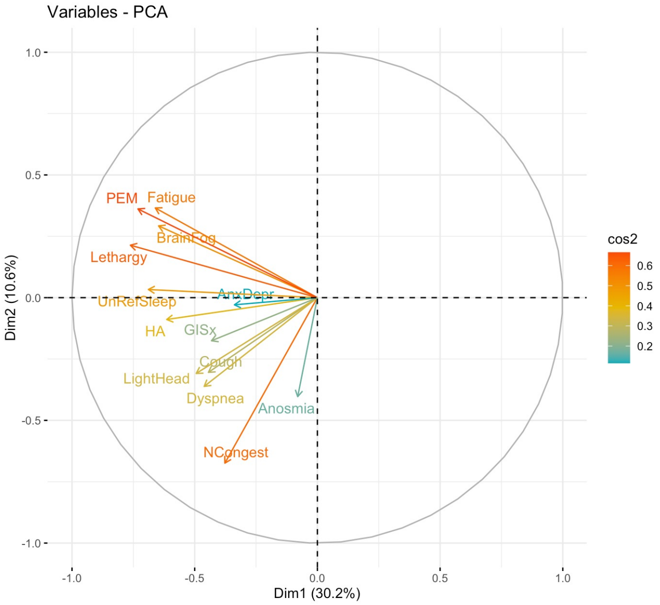

#### Total Symptoms, and distribution by sex

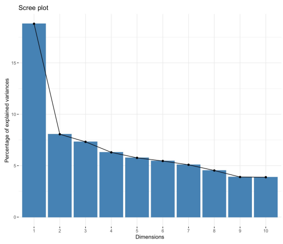

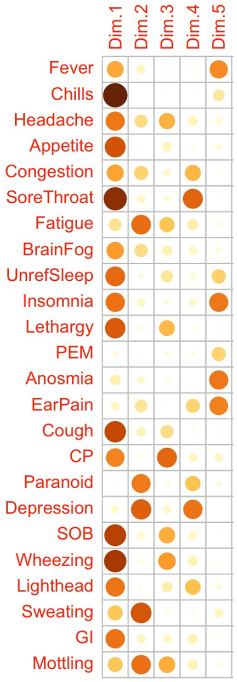

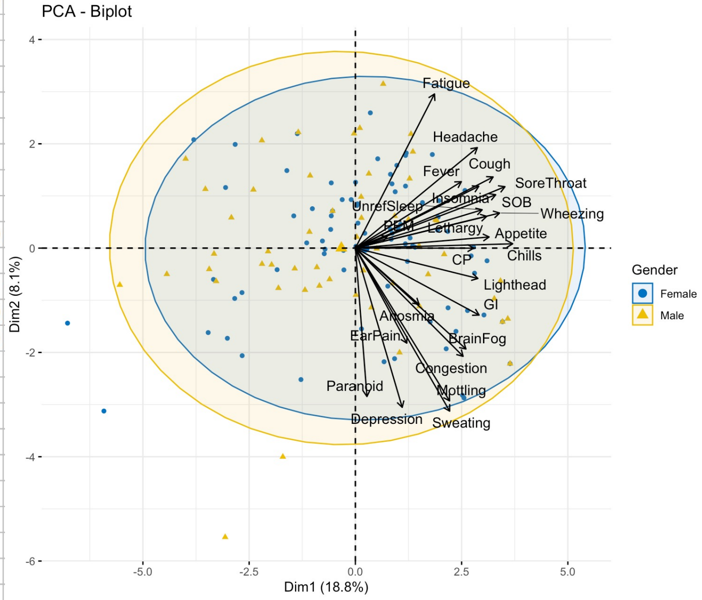

Common symptoms male:

| | eigenvalue | variance.percent | cumulative.variance.percent |
|---|---|---|---|
| Dim.1 | 3.1318023 | 24.090787 | 24.09079 |
| Dim.2 | 1.7423991 | 13.403070 | 37.49386 |
| Dim.3 | 1.4808081 | 11.390832 | 48.88469 |
| Dim.4 | 1.2178200 | 9.367846 | 58.25253 |
| Dim.5 | 1.0799056 | 8.306966 | 66.55950 |
| Dim.6 | 1.0063531 | 7.741177 | 74.30068 |
| Dim.7 | 0.7773718 | 5.979783 | 80.28046 |
| Dim.8 | 0.6786311 | 5.220239 | 85.50070 |
| Dim.9 | 0.5662909 | 4.356084 | 89.85678 |
| Dim.10 | 0.4300200 | 3.307846 | 93.16463 |
| Dim.11 | 0.3619187 | 2.783990 | 95.94862 |
| Dim.12 | 0.2912270 | 2.240208 | 98.18883 |
| Dim.13 | 0.2354524 | 1.811172 | 100.00000 |

Common symptoms female:

| | eigenvalue | variance.percent | cumulative.variance.percent |
|---|---|---|---|
| Dim.1 | 4.6530112 | 35.792393 | 35.79239 |
| Dim.2 | 1.3126076 | 10.096982 | 45.88938 |
| Dim.3 | 1.1123348 | 8.556422 | 54.44580 |
| Dim.4 | 1.0321811 | 7.939854 | 62.38565 |
| Dim.5 | 0.9694872 | 7.457594 | 69.84325 |
| Dim.6 | 0.8592313 | 6.609471 | 76.45272 |
| Dim.7 | 0.7104871 | 5.465285 | 81.91800 |
| Dim.8 | 0.6237441 | 4.798032 | 86.71603 |
| Dim.9 | 0.4749512 | 3.653471 | 90.36950 |
| Dim.10 | 0.4092251 | 3.147885 | 93.51739 |
| Dim.11 | 0.3378931 | 2.599177 | 96.11657 |
| Dim.12 | 0.2790088 | 2.146221 | 98.26279 |
| Dim.13 | 0.2258376 | 1.737212 | 100.00000 |

Common symptoms total:

| | eigenvalue | variance.percent | cumulative.variance.percent |
|---|---|---|---|
| Dim.1 | 3.9263275 | 30.202520 | 30.20252 |
| Dim.2 | 1.3727348 | 10.559499 | 40.76202 |
| Dim.3 | 1.1229366 | 8.637974 | 49.39999 |
| Dim.4 | 1.0777255 | 8.290196 | 57.69019 |
| Dim.5 | 1.0180607 | 7.831236 | 65.52142 |
| Dim.6 | 0.9121950 | 7.016885 | 72.53831 |
| Dim.7 | 0.7759026 | 5.968481 | 78.50679 |
| Dim.8 | 0.6279203 | 4.830156 | 83.33695 |
| Dim.9 | 0.5602774 | 4.309826 | 87.64677 |
| Dim.10 | 0.4973165 | 3.825512 | 91.47228 |
| Dim.11 | 0.4072518 | 3.132706 | 94.60499 |
| Dim.12 | 0.3895180 | 2.996292 | 97.60128 |
| Dim.13 | 0.3118331 | 2.398717 | 100.00000 |

Eigenvalues 12 domains (Top total, Middle female and low male)

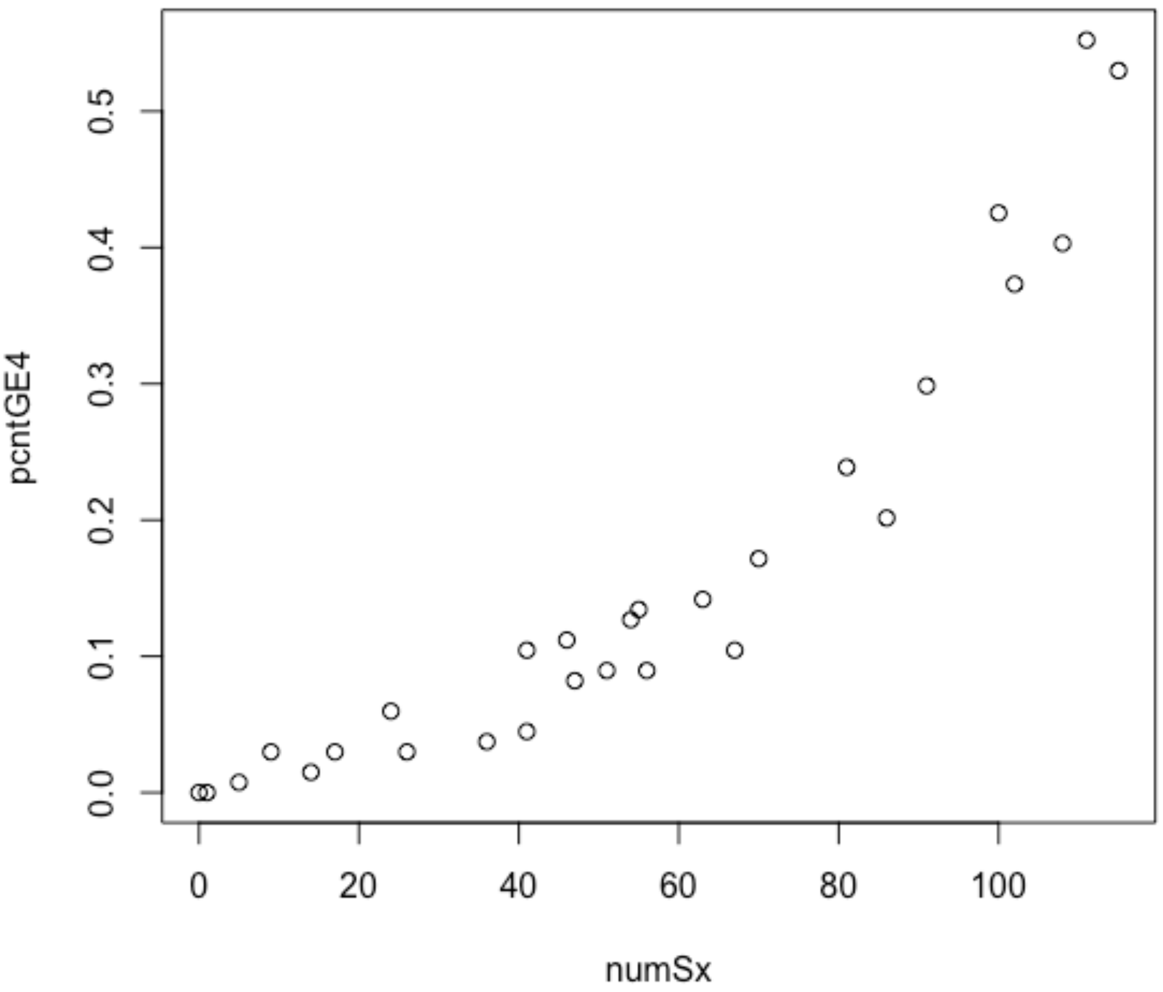

Parson Correlation Coefficient (PCC) frequency of 29 symptoms and severity (Likert scale 4 and 5).
